# Methylation-driven Cancer Genes and Methylation Profiling in Glioma: A Comparative Study between East Asian and non-Hispanic White Populations

**DOI:** 10.64898/2026.08.14.26360452

**Authors:** Louise Newman, Niall Dunne, Vinton WT Cheng, Archana Sharma-Oates

**Author notes:** Correspondence to Archana Sharma-Oates.

## Abstract

Global incidence and outcomes of glioma have been found to vary significantly by region, however research into the disease continues to lack diversity. Here we investigated epigenetic patterns in glioma subtypes from cohorts collected from China and the USA.

We retrospectively analysed the Chinese Glioma Genome Atlas (CGGA) and The Cancer Genome Atlas (TCGA) datasets following reclassification of glioma subtypes based on the WHO 2021 central nervous system (CNS) tumour classification. We used DNA methylation and transcriptomics data to identify methylation-driven cancer genes in the CGGA cohort, assessed their prognostic value and compared against the non-Hispanic White cohort in the TCGA database to consider ethnic influence. Furthermore, we used machine learning classification and clustering techniques to identify methylation patterns in glioma subgroups.

Here, we showed that DNA methylation profiles of CGGA glioblastomas have a methylation signature more similar to TCGA high-grade astrocytomas: 36.7% of CGGA glioblastomas (n=49) were identified as high-grade astrocytomas using classification modelling. Assessment of survival revealed that CGGA glioblastoma patients had a significantly better survival rate than non-Hispanic White glioblastoma patients (p = 0.037). Four key methylation-driven genes were identified in the CGGA glioblastoma samples: GLDN, PRKDC, S100A1 and NCAPH. Hypermethylation of GLDN significantly suppressed gene expression in all glioma subtypes in only the East Asian cohort; a gene that has not been previously described as a driver in gliomas.

Together these data suggest alternative epigenetic mechanisms occurring in glioma subtypes of different ethnic populations, which is important for our understanding of glioma and strategies for personalized treatment.

## Background

Gliomas are tumours of glial cell lineage, most often arising from astrocytes, oligodendrocytes or ependymal cells, and are the most common malignant brain tumour. Incidence of glioma has been found to vary globally, with regions in Asia amongst the areas with lowest incidence for various glioma subtypes^1^. In a study investigating glioma survival rates in patients of various ethnicities within the US, participants of European ancestry were found to have a higher incidence of glioma as well as experiencing lower survival rates^2,3^. Reporting of glioma outcomes globally are confounded by inequitable availability of diagnostic tools and treatment, however there are several environmental hazards and lifestyle choices that have been shown to affect the risk of gliomas, which could contribute to global disparities in risk and survival^4–8^.

Epigenetics is the study of heritable changes to gene expression that do not alter the DNA sequence, influenced by environmental and lifestyle factors. DNA methylation is one such epigenetic modification, causing transcriptomic silencing through the addition of methyl groups in the DNA sequence. Epigenetic-targeting agents have been identified as a promising therapeutic strategy in the treatment of gliomas; to improve response to existing therapies as well as having the potential to offer personalised disease treatment^9^. However, for epigenetics to play an effective part of glioma treatment for all patients, it is crucial that typically underrepresented groups are included in this research. A recent investigation into glioma involving omics data revealed a substantial lack of reporting of ethnicity information and in those studies where ethnicity was reported, there was a considerable underrepresentation of many minority ethnic groups^10^.

Accordingly, in this study we investigated DNA methylation profiles in glioma subtypes in participants from the Chinese Glioma Genome Atlas (CGGA) and compared the findings to participants from The Cancer Genome Atlas (TCGA) who had self-reported their ethnicity as non-Hispanic White. This cohort was chosen for comparison as it is the largest ethnic population on the TCGA archive. The CGGA database is a comprehensive resource which includes glioma tumour samples collected from multiple centres across Northern and Eastern regions of China^11^. We hypothesized that population-specific epigenetic differences contribute to variation in glioma survival and sought to determine whether distinct DNA methylation signatures could be attributed to glioma from different ethnic populations. Furthermore, we sought to investigate methylation-driven cancer genes in the East Asian cohort, to identify prognostic biomarkers and epigenetic drivers that may be overlooked in studies with limited diversity.

## Results

### High-grade astrocytomas and glioblastomas are associated with significantly different survival outcomes between ethnic groups that are not attributed to epigenetic age acceleration

Participants with gliomas classified as low-grade astrocytomas (LGA) showed no significant differences in survival rates between CGGA and TCGA cohorts (median overall survival (mOS; months) = not reached (CGGA) vs 145 (TCGA); p = 0.8) (Fig 1a). However, in participants with high-grade astrocytomas (HGA), survival was significantly poorer in those from the CGGA cohort (mOS = 17.5 (CGGA) vs 62.0 (TCGA); *p* = 0.033) (Fig 1b). Conversely, in participants with glioblastoma (GBM), the survival of the CGGA cohort was significantly better compared to those from the TCGA cohort (mOS = 18.5 (CGGA) vs 14.0 (TCGA); *p* = 0.037) (Fig 1c).

**Figure 1.**
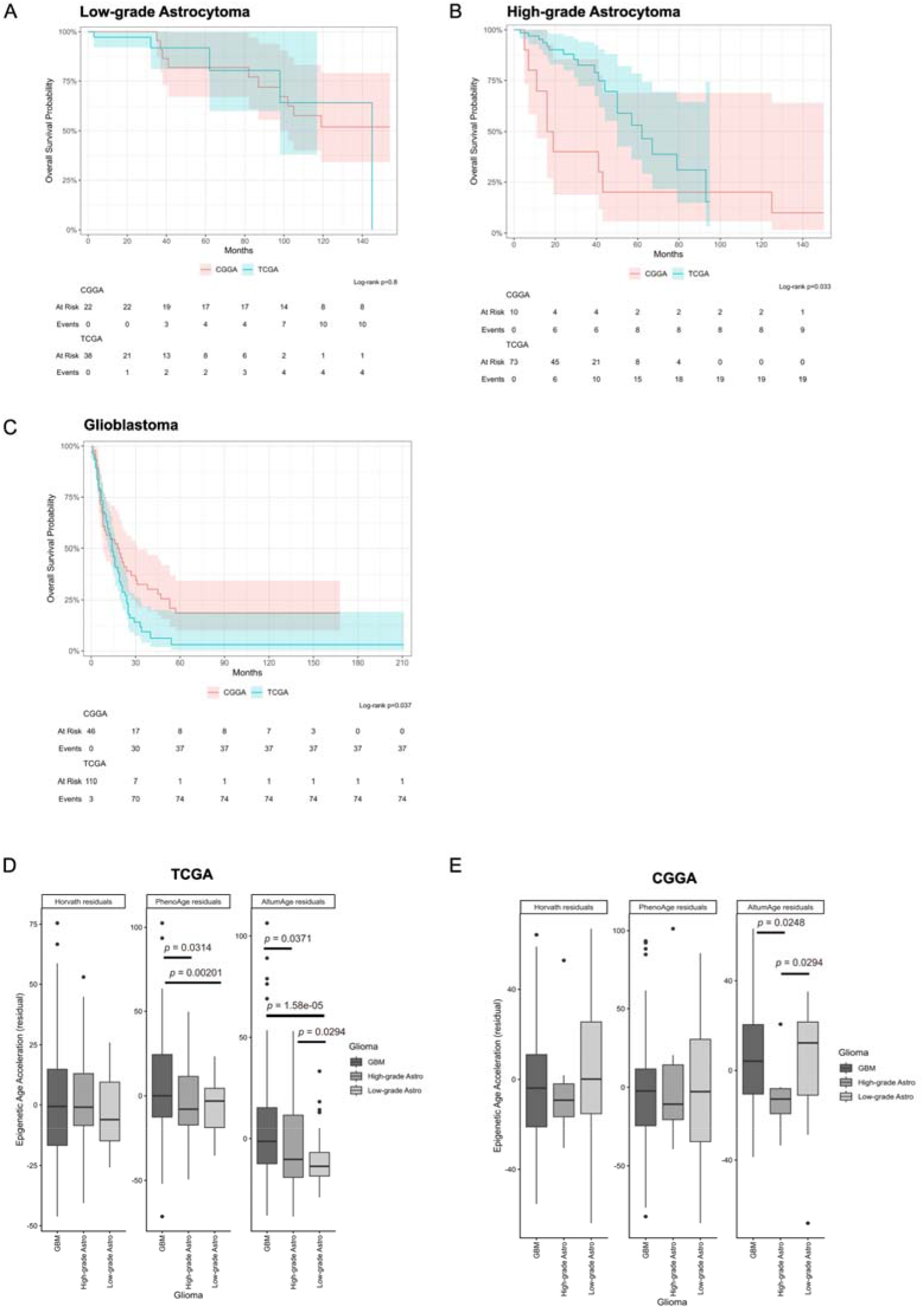
Kaplan-Meier survival analysis and epigenetic age acceleration analysis of CGGA and TCGA datasets. A-C. Kaplan-Meier survival analysis plots for CGGA (red) and TCGA (blue) participants, separated by glioma type: (A) Low-grade astrocytomas (B) high-grade astrocytomas, (C) glioblastoma. D-E. Box and whisker plots of epigenetic age acceleration, measured using residuals, as estimated using epigenetic clocks Horvath (2013), PhenoAge and AltumAge in TCGA (D) and CGGA (E) cohorts. Statistical significance was assessed within each cohort, comparing different glioma types, using two-tailed *t*-tests and *p*-values reported where significant.

The three epigenetic clocks (Horvath 2013, PhenoAge and AltumAge) used in the study were chosen due to their compatibility with both 27K and 450K methylation arrays. Gliomas typically display vast acceleration and deceleration of biological age compared to chronological age, and this phenomenon was replicated in the samples used in this study, spanning a range of −64 to +144 years of biological deviation, consistent with previous reports (Supplementary Fig 1b-d)^12,13^. Despite this, when comparing epigenetic age deviation from chronological age within each cohort, significant differences were found between glioma types (Fig 1d, e). In the TCGA cohort, two clocks reported significantly increased epigenetic age acceleration of GBM samples, typically associated with the poorest survival outcomes, compared to both other glioma types (PhenoAge, mean epigenetic age residual (mEAR): GBM 5.29 29.78, HGA −3.20 23.02 (*p* = 0.031), LGA −6.43 14.78 (*p* = 0.0020); AltumAge, mEAR: GBM 3.98 25.37, HGA −3.49 20.50 (*p* = 0.037), LGA −10.64 13.39 (*p* = 1.6e-05)) (Fig 1d). The AltumAge clock also found a significant acceleration in HGAs compared to LGAs (*p* = 0.030), suggesting a relationship between increasing glioma grade and epigenetic age acceleration that was not observed in the CGGA cohort (Fig 1e). Paradoxically, using the AltumAge clock on the CGGA data, a deceleration was observed between LGA and HGA samples (AltumAge mEAR: HGA −12.57 13.52, LGA 4.85 23.74 (*p* = 0.029)) (Fig 1e).

When comparing glioma types between cohorts, a significant difference was only found in one clock (AltumAge) in LGA samples (AltumAge mEAR: CGGA LGA 4.85 23.74, TCGA LGA −10.64 13.39 (*p* = 0.0047); Supplementary table 4).

### DNA methylation signature of glioblastoma in CGGA cohort is comparable to high-grade astrocytomas in TCGA cohort

Feature selection for the TCGA-trained random forest model was determined by selecting significantly differentially methylated CpG probes with large log fold changes (LFC) between glioma subtypes (GBM vs HGA: LFC > 4, 133 probes, GBM vs LGA: LFC > 4, 154 probes, HGA vs LGA: LFC > 1.2, 97 probes). Seventy percent of the TCGA cohort, with 265 unique CpG probes, was used to train the random forest model and using the test TCGA cohort (30%) it was found to perform well at classifying GBM and HGA (overall accuracy: 0.923, sensitivity (GBM) = 1.0, sensitivity (HGA) = 1.0). However, performance was less accurate when classifying LGA (sensitivity (LGA) = 0.545; Supplementary Fig 2c). During feature selection, the differentially expressed probes had smaller log fold changes between low- and high-grade astrocytomas, compared to other combinations of the three glioma subtypes, which may explain the poorer performance. Consequently, we decided to focus on GBM and HGA only, to ensure a fair comparison of the two ethnic cohorts. In the next TCGA-trained classifier, using the same CpG probes for GBM and HGA samples, the performance on TCGA test data had a strong overall accuracy (0.982; Fig 2a, Supplementary table 5). For the CGGA cohort, 36.7% of the GBM samples were misclassified as HGA (Fig 2b). CGGA high-grade astrocytomas appeared to have a very similar methylation profile to TCGA high-grade astrocytomas when considering these particular probes (90.9% of CGGA HGAs were correctly classified; Fig 2b).

**Figure 2.**
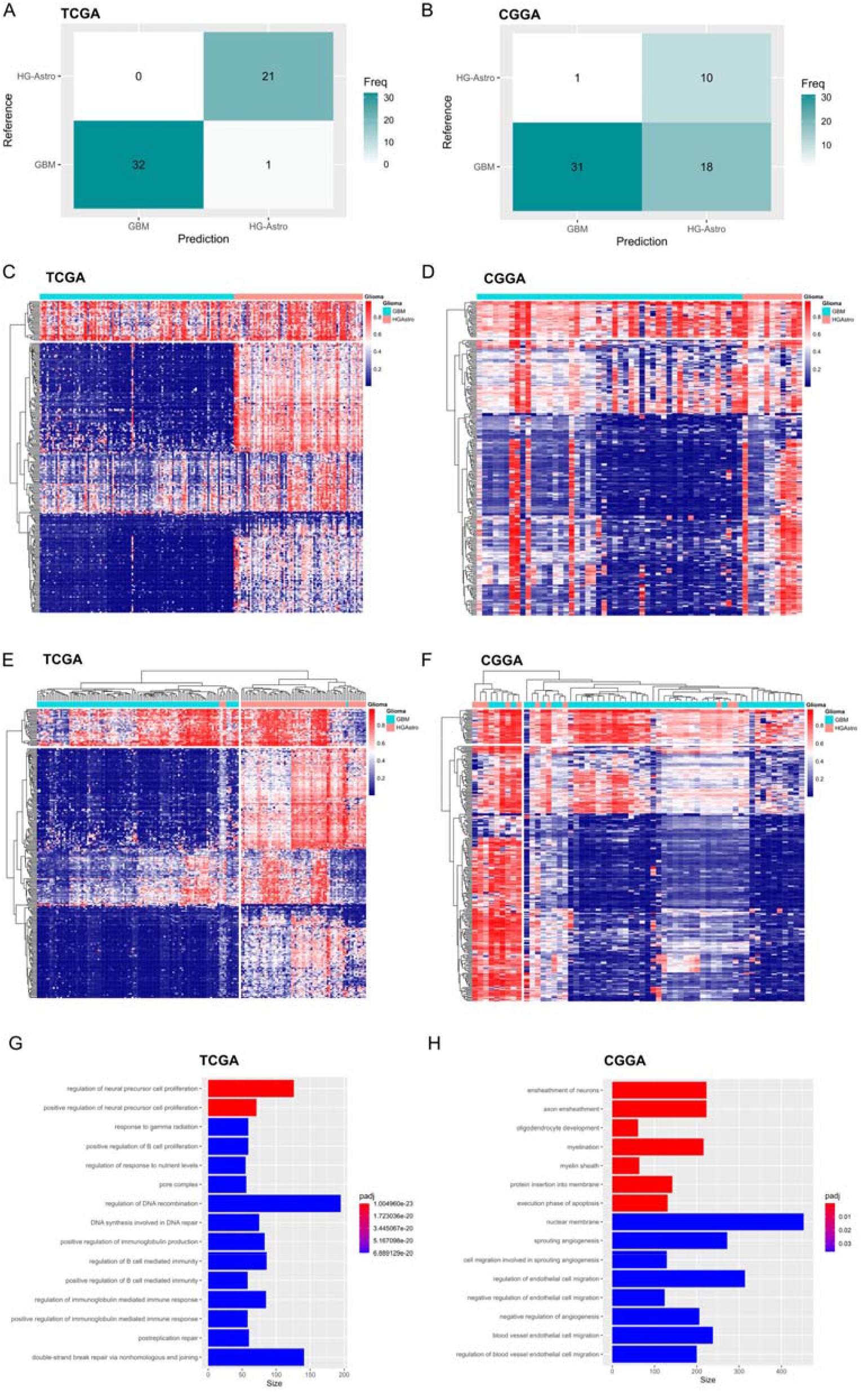
Classification and clustering of high-grade astrocytomas and glioblastomas using DNA methylation profiles. A. Confusion matrix showing classification performance of random forest modelling, trained on TCGA glioblastoma and high-grade astrocytoma samples only, using 265 CpG probes and tested on TCGA test data. B. Confusion matrix of the TCGA-trained model performance on CGGA data. C-D. DNA methylation heatmaps showing beta values, with hierarchical clustering of the 265 CpG probes used in the RF model, for TCGA (C) and CGGA (D). Blue indicates hypomethylation and red indicates hypermethylation. E-F. DNA methylation heatmaps as in (C-D), with hierarchical clustering of samples for TCGA (E) and CGGA (F). G-H. Gene ontology enrichment analysis of differentially methylated probes between glioblastoma and high-grade astrocytomas in TCGA (G) and CGGA (H). Statistical significance was determined using a logistic regression model in methylglm, with Benjamini-Hochberg correction for multiple testing.

The beta values of the 265 selected CpG probes showed a distinct difference in the TCGA cohort of GBM and HGA samples, with the former appearing to have many more hypomethylated probes compared to the latter (TCGA mean beta value: GBM 0.200 0.053, HGA 0.501 0.091 (*p* = 2.2e-16), Fig 2c). Conversely, the CGGA GBMs had a less consistent methylation profile for the same probes, although the mean beta values of the GBM and HGA gliomas were still significantly different from each other (CGGA mean beta value: GBM 0.325 0.144, HGA 0.512 0.156 (*p* = 0.00032), Fig 2d). Under hierarchical clustering, the TCGA samples were almost exclusively clustered according to the glioma subtype, however in the CGGA cohort, the gliomas subtypes were unevenly distributed.

To investigate further, differential analysis was conducted on CGGA and TCGA DNA methylation data separately. The significantly differentially methylated probes between GBM and HGA (>2.5 log fold change relative increase or decrease) underwent gene ontology (GO) enrichment analysis. Notably, there was no overlap of *any* significant GO terms between the two cohorts (Fig 2g, h).

### Identification of methylation-driven cancer genes in the CGGA cohort

Analysis of differentially methylated genes and gene expression identified a total of 15 MDCGs in gliomas compared to control samples (Table 1). No overlap in GO terms was found in MDCGs between glioma subtypes. GO pathway analysis using the glioblastoma MDCGs identified seven significantly enriched GO terms, involving genes related to cell-cell adhesion, channel clustering and neuronal maturation, which are processes that have been previously associated with tumour infiltration and proliferation (Fig 3a)^14–16^.

**Figure 3.**
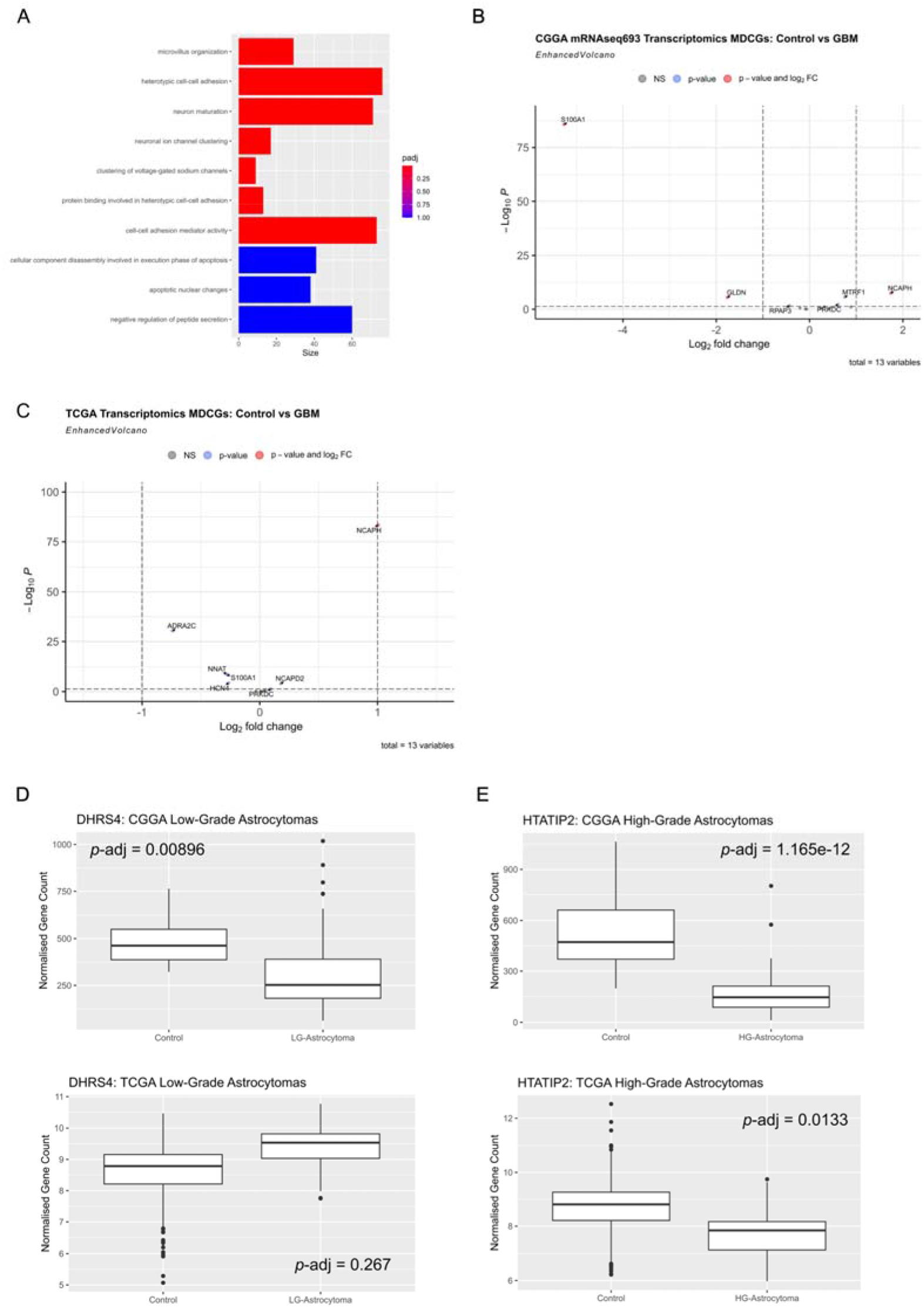
Methylation-driven cancer genes: pathway and transcriptomic analysis in CGGA gliomas. A. Enrichment analysis of gene ontology terms conducted on adjusted p-values of MDCGs following differential analysis of M-values of the 13 identified MDCGs (corresponding to 23 CpG probes) between control and glioblastoma samples from the CGGA cohort. B. Volcano plot of gene expression changes in the 13 MDCGs in another CGGA glioblastoma cohort (mRNAseq693) following differential gene expression analysis of all available genes compared to CGGA controls. Significant gene expression changes were determined by the Wald test in DESeq2 with Benjamini-Hochberg correction for multiple testing, and are represented by blue circles. Red circles indicate statistical significance with a log fold change of > ±1. C. Volcano plot of gene expression changes of the 13 MDCGs in the TCGA glioblastoma dataset following differential gene expression analysis of all available genes compared to TCGA-GTEx controls. D. Box and whisker plot of normalized gene counts for the DHRS4 gene in low-grade astrocytoma samples and controls (CGGA: left, TCGA: right) following differential gene analysis with p-adjusted significance values. E. Box and whisker plot of normalized gene counts for the HTATIP2 gene in low-grade astrocytoma samples and controls (CGGA: left, TCGA: right) following differential gene analysis with p-adjusted significance values. Statistical significance was determined using the Wald test in DESeq2 with Benjamini-Hochberg correction for multiple testing.

**Table 1.**
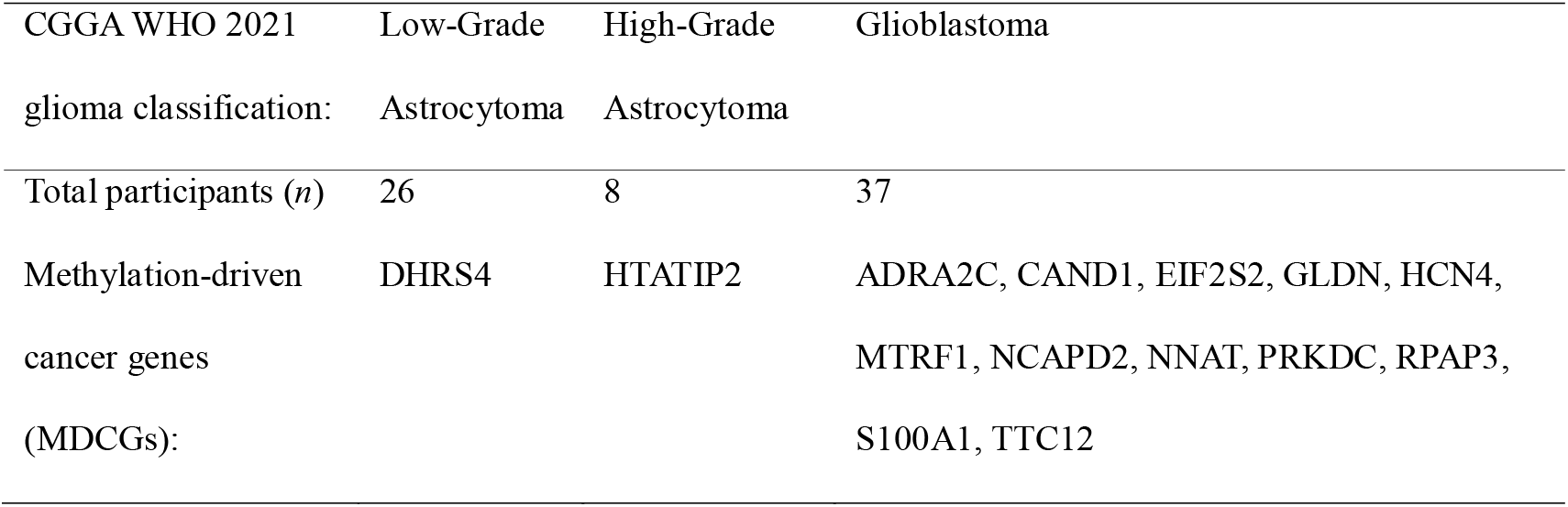
Methylation-driven cancer genes (MDCGs) identified in CGGA glioma samples. List of MDCGs in the CGGA dataset determined by comparing DNA methylation data from each glioma subtype (low-grade astrocytoma, high-grade astrocytoma and glioblastoma) to control data.

Four novel MDCGs identified in GBM samples have not been directly associated with glioma previously to our knowledge: CAND1, GLDN, HCN4 and MTRF1. Of the 13 MDCGs associated with GBM, only 6 were found to have significantly different gene expression compared to controls in a second CGGA transcriptomics dataset, and 7 were found to be differentially expressed in the TCGA dataset (Fig 3b, c). Only three overlapped between CGGA and TCGA: NCAPH, S100A1 and PRKDC; with the direction of change of these genes found to be consistent between the cohorts.

In the LGA and HGA samples, gene expression changes of both subtype-associated MDCGs, DHRS4 and HTATIP2 respectively, were validated in the second CGGA dataset, but only HTATIP2 was also found to be significantly differentially expressed in the TCGA cohort (Fig 3d, e).

### Hypomethylation of PRKDC is associated with worse survival outcomes in the CGGA cohort

To investigate MDCGs from the CGGA glioblastoma cohort further, we assessed the methylation profiles of the CpG probes from the six validated genes: GLDN, MTFR1, PRKDC, S100A1, RPAP3 and NCAPH. Further investigation using gene expression data from the CGGA validation cohort revealed that RPAP3 was not methylation-driven, since the significant gene expression changes were not in the opposite direction to methylation changes (Fig 3b, 4a, Supplementary Fig 3). By visualizing methylation data by individual probes, it was noted that any changes in beta values of the MTRF1 gene, measured at either CpG site, were not considerably different to controls. Therefore, it was decided that further analysis would be conducted on GLDN, PRKDC, S100A1 and NCAPH.

**Figure 4.**
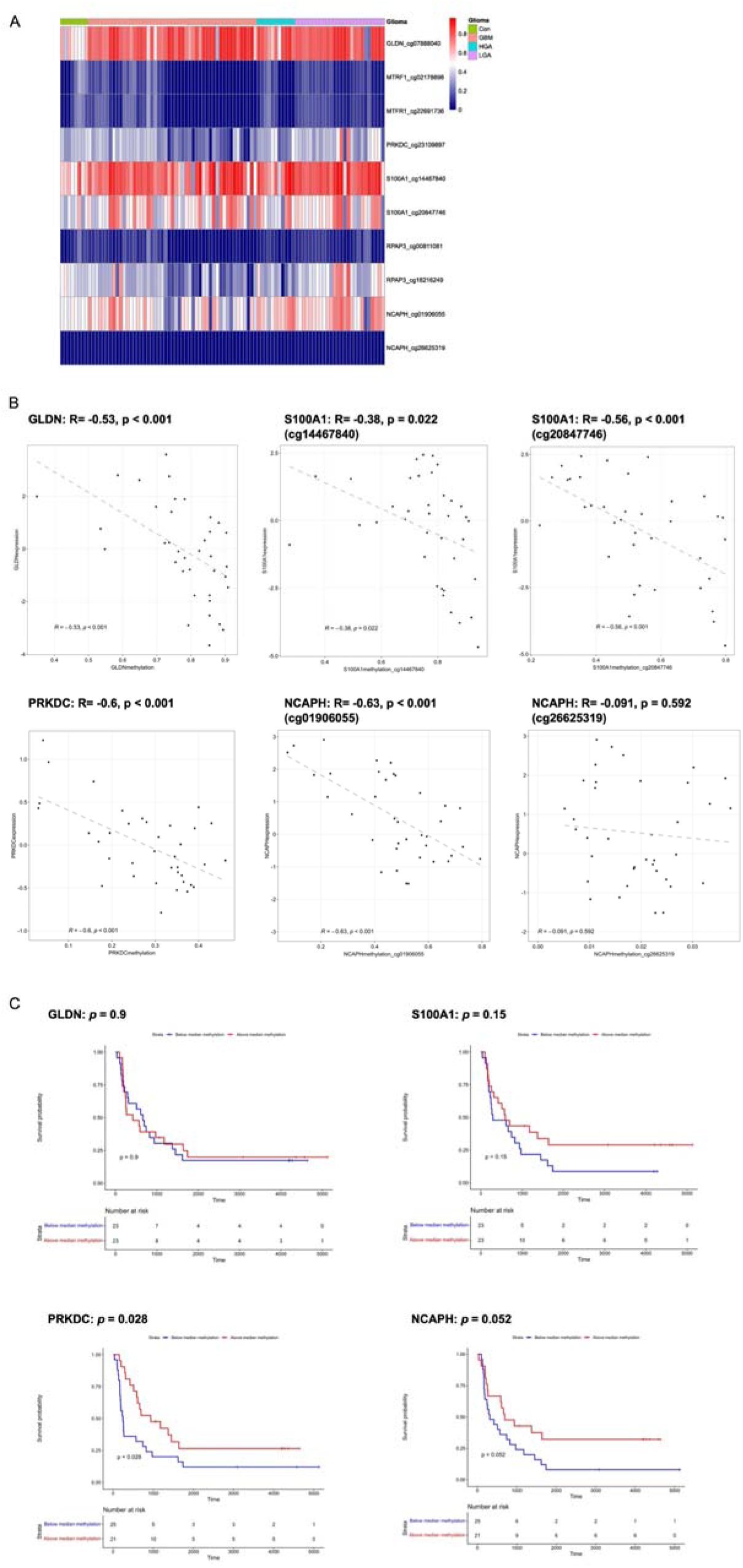
Glioblastoma MDCGs: correlation of DNA methylation probes with transcriptomics data and prognostic value. A. DNA methylation heatmaps showing beta values (ranging from 0-1, blue to red) for each CpG probe of the six validated MDCGs identified from the CGGA glioblastoma dataset. B. Scatter plots showing correlations between the methylation and expression levels of four validated glioblastoma MDCGs of interest. Genes S100A1 and NCAPH had two CpG probes available in the 27K array data. This data uses CGGA glioblastoma samples only, and the methyl_159 and mRNA-array_301datasets. Correlation coefficient was determined using Pearson’s correlation. C. Kaplan-Meier survival analysis plots for CGGA glioblastoma samples separated into two groups according to methylation levels of each gene. Samples with DNA methylation beta values higher than the median are labelled in red, and those with beta values below the median are labelled in blue. Statistical significance was determined using the log-rank test.

Next, we performed multi-omic correlation to investigate the relationship between DNA methylation and gene expression in glioma tumours (Fig 4b). Using the CGGA GBM dataset, we confirmed that all four genes had a significant inverse correlation between methylation and gene expression (Fig 4b). Where multiple probes mapped to the same gene, we identified which of these had the strongest effect on gene expression, thereby identifying probe-specific methylation-driven genes (Fig 4b). We then conducted univariate survival analysis on this cohort, by separating participants into high and low methylation groups (high = above median beta value, low = below median beta value) and assessing the survival curves (Fig 4c). A significant difference was found in the effect of PRKDC methylation, whereby a below-median methylation level was associated with poorer survival outcomes.

### Methylation of PRKDC at the same CpG site did not correlate with gene expression or associate with patient survival in the TCGA dataset

To investigate whether this finding could be replicated in the TCGA cohort, we assessed the same CpG site and found a wide range of beta values in the glioblastoma samples, which did not correlate with gene expression of PRKDC (Fig 5a, b). Unlike the CGGA cohort, no differences were found in patient survival curves when samples were stratified by PRKDC methylation beta values (Supplementary Fig 4). Our ability to assess methylation of a particular gene is limited by the CpG sites at which methylation is determined. The CGGA database uses 27K array technology, however the TCGA database has both 27K and the much more extensive 450K data available. We therefore used the glioblastoma participants for which we had 450K array data (*n* = 72) to investigate other PRKDC CpG sites, and found that in this dataset these were overwhelmingly hypermethylated (Fig 5c). Of the 20 CpG sites available in the 450K dataset, none were significantly correlated with gene expression of PRKDC, however methylation at just one probe did reveal a significant difference in survival when samples were stratified by PRKDC methylation beta values (Supplementary table 6, Fig 5d). Interestingly, in this cohort, the higher methylation levels were associated with poorer patient survival. Due to technical limitations, we were unable to assess this same probe in the CGGA dataset.

**Figure 5.**
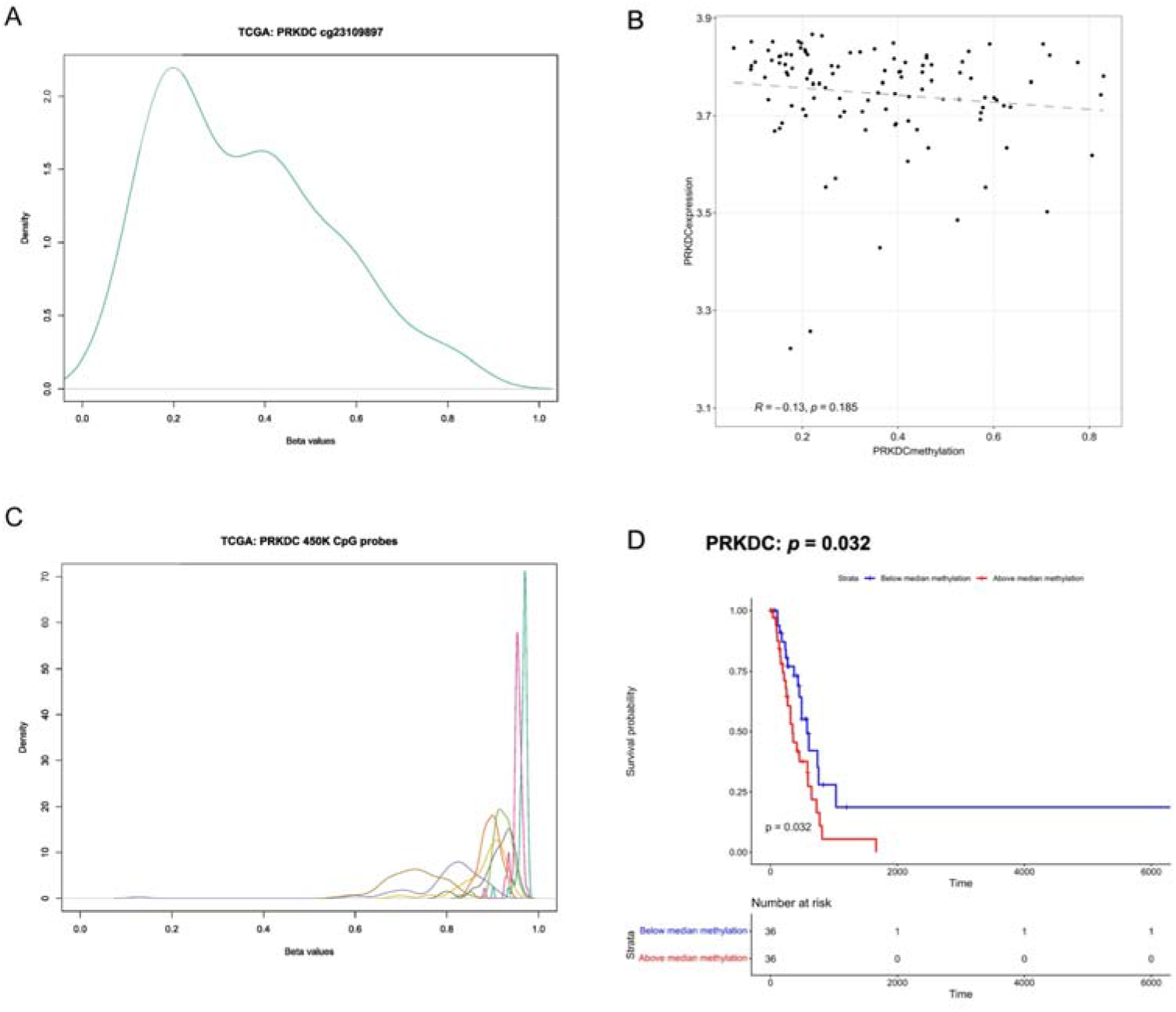
Analysis of DNA methylation and gene expression of the PRKDC gene in the TCGA cohort. A. Beta density plot of the PRKDC CpG probe beta values (cg23109897) available in 27K array data for glioblastoma samples from the TCGA cohort. This data has undergone normalization to correct for differences in data obtained from the 27K and 450K DNA methylation arrays. B. Scatter plot showing correlation between DNA methylation and gene expression of the PRKDC gene in glioblastoma samples from the TCGA cohort. This data uses beta values from the DNA methylation probe cg23109897, normalized as in (A), and the gene expression data is normalized VCT counts. Correlation coefficient was determined using Pearson’s correlation. C. Beta density plot of the PRKDC CpG probes available in the 450K array data, using glioblastoma samples from the TCGA cohort obtained using 450K array technology. These data were normalized to correct for batch differences in the glioblastoma samples (WHO 2021 classification) obtained from the TCGA-LGG and TCGA-GBM datasets. CpG probes are coloured to allow for easier visualisation. D. Kaplan-Meier survival analysis plot of TCGA glioblastoma samples separated by PRKDC methylation beta value for the 450K CpG probe cg14189875. Samples with DNA methylation beta values higher than the median are labelled in red, and those with beta values below the median are labelled in blue. Statistical significance was determined using the log-rank test.

### Hypermethylation causes suppression of gliomedin gene expression in the CGGA cohort but not in TCGA samples

The identification of GLDN as a MDCG in the CGGA glioblastoma dataset was of particular interest, since this is a gene that has not been previously associated with glioblastoma. The DNA methylation profiles of all glioma subtypes revealed a hypermethylation of GLDN compared to controls and gene expression of GLDN was significantly reduced in all glioma tumours (Fig 4a, Fig 6a). However, analysis of the TCGA dataset found no significant differences in GLDN gene expression (Fig 6b). Whilst GLDN has been identified as a cancer driver, analysis of both DNA methylation and gene expression did not reveal it to be prognostic in glioblastoma samples (Fig 4c, Fig 6c). Unsupervised clustering of GLDN methylation values revealed three methylation subgroups, however these did not correspond with glioma subtype classification (Supplementary Fig 5, Fig 6d).

**Figure 6.**
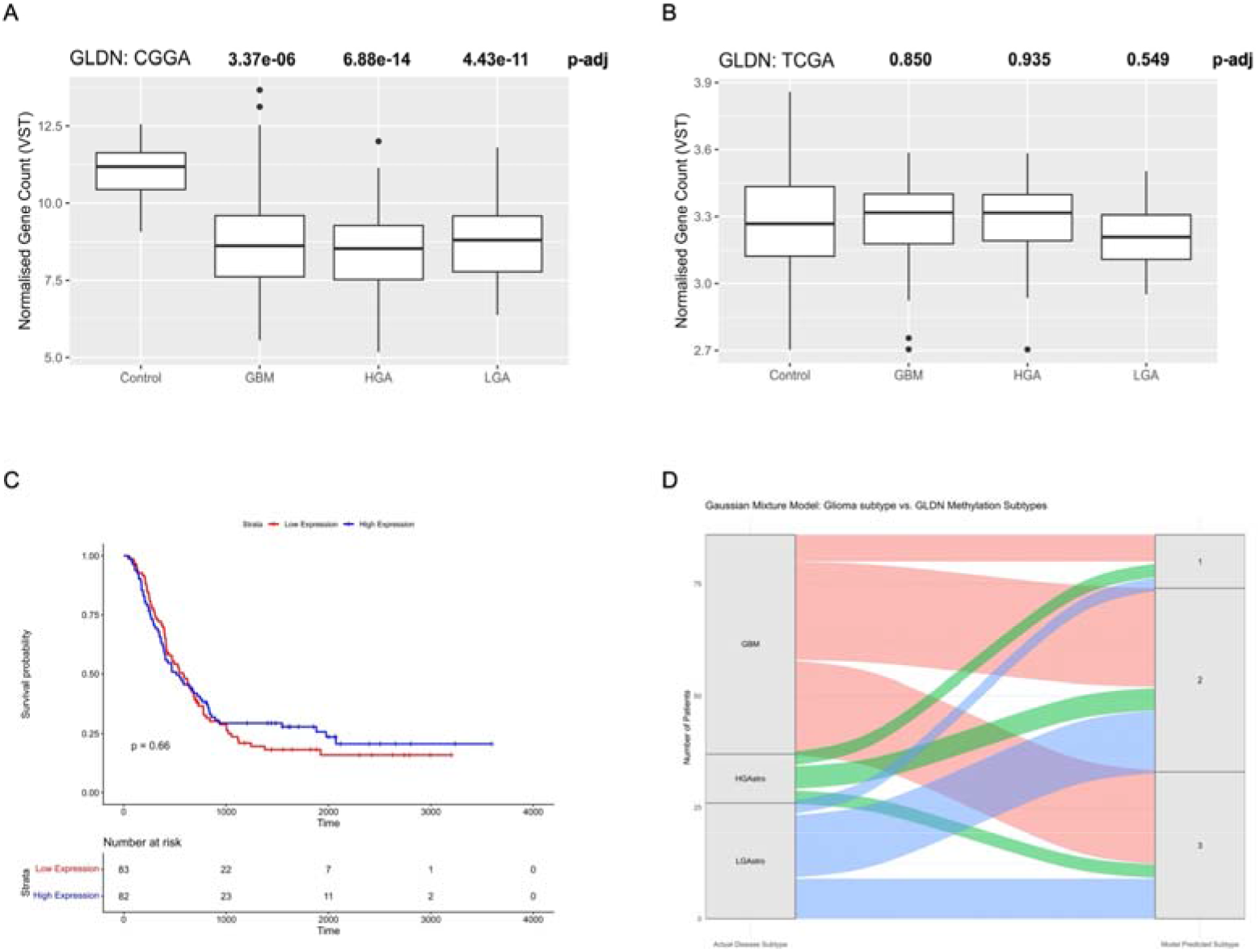
Analysis of gene expression and DNA methylation of GLDN in CGGA and TCGA cohorts. A. Gene expression analysis of GLDN in CGGA gliomas (GBM = glioblastoma, HGA = high-grade astrocytomas, LGA = low-grade astrocytomas) using the mRNAseq_693 dataset. Box plots show normalized VST gene counts and statistical significance was determined following differential gene expression analysis between glioma and control samples, using the Wald test in DESeq2 with Benjamini-Hochberg correction for multiple testing. B. Gene expression analysis of GLDN in gliomas from the TCGA cohort, as in (A). C. Kaplan-Meier survival analysis of CGGA glioblastomas separated by GLDN gene expression levels. Samples with gene expression levels higher than the median are labelled in red, and those with gene expression below the median are labelled in blue. Statistical significance was determined using the log-rank test. D. Alluvial diagram generated using CGGA gliomas (GBM = glioblastoma, HGAstro = high-grade astrocytoma, LGAstro = low-grade astrocytoma) showing the actual glioma subtype (left) and the predicted epigenetic subtype (right) based on a Gaussian mixture model using GLDN DNA methylation data.

## Discussion

The Cancer Genome Atlas is considered one of the most cited cancer genomics databases, and the CGGA database is frequently used as a validation cohort for studies analysing the TCGA-GBM and TCGA-LGG datasets. However, it is known that DNA methylation disparities exist in both healthy and cancerous tissues; for example, as a result of methylation quantitative trait loci (meQTL) that are present in particular populations^17,18^. Ethnic variation in DNA methylation profiles has been identified in many diseases and several cancer tumours, including prostate and breast cancer^19–25^. We hypothesised that population-specific epigenetic differences between these two cohorts may affect glioma tumour biology and disease prognosis.

In this study we used the CGGA dataset and updated tumour definitions using the 2021 WHO CNS tumour classification to identify and validate MDCGs. Of the validated glioblastoma MDCGs, only one gene, PRKDC, was also differentially expressed in the TCGA dataset. However, this did not appear to be caused by hypomethylation at the same CpG site, located within the promoter region, as in the CGGA dataset. PRKDC has a role in the repair of DNA double-strand breaks, and in gliomas its overexpression has been recently linked to reduced survival and treatment resistance, however this has not previously been shown to be methylation-driven^26^. Since we were able to consider more CpG sites in some samples of the TCGA data, hypermethylation of the PRKDC gene at a different CpG site, within the gene body, significantly associated with poorer patient outcomes, suggesting an alternative mechanism to that seen in the CGGA dataset: hypomethylation within the promoter gene increases gene transcription, however hypermethylation within the gene body can activate gene expression^27^. Both processes can drive tumour survival, but the mechanisms appear to be inverted in these two cohorts and therapeutically targeting this would require opposing epigenetic treatment strategies.

By focussing on the CGGA cohort, we were also able to implicate GLDN as a driver gene in glioma. Hypermethylation of GLDN caused a significant reduction in gene expression in all glioma subtypes within in the CGGA dataset, a finding not replicated in the TCGA samples. GLDN has a role in the formation of nodes of Ranvier, and in cancer it has been identified as both a tumour suppressor gene and an oncogene^28–30^. Neither GLDN methylation or expression were prognostic in our analysis, so it is unclear what the effect of reduced GLDN expression has on glioma biology and clinical outcomes.

DNA methylation patterns are used for the profiling and classification of glioma tumours, in combination with histological imaging, known as the Heidelberg CNS Tumor Methylation Classifier^31^. Recently the classifier has been expanded to include data from more global regions, however the diversity of participants still does not reflect that of glioma patients worldwide^32^. Unfortunately, we were unable to use the Heidelberg Classifier on the CGGA dataset as it requires data from more CpG sites that are obtained using newer technology. As a result, in this study we generated our own random-forest classifier using TCGA data and used it to compare the methylation profile of glioma subtypes between our two cohorts. Firstly, we demonstrated that DNA methylation patterns of high-grade astrocytomas were comparable between these two cohorts, suggesting similar epigenetic features for this particular tumour type at the methylation sites considered. Secondly, however, this was not the case for glioblastoma samples: more than a third of the CGGA glioblastoma tumours, defined as lacking isocitrate dehydrogenase (IDH) mutations, were more akin to those of high-grade astrocytomas of the TCGA data, which possess an IDH mutation. Interestingly, within the glioblastoma patients, survival outcomes for those from the CGGA cohort were significantly better than those from the TCGA cohort. The heterogeneity of the CGGA glioblastoma samples is suggestive of additional subtypes, a concept that is well established in glioblastoma studies, which may occur as a result of tumours that progress from a lower-grade or reflect the complex tumour microenvironment^33–35^. It could be that certain epigenetic influences increase the prevalence of particular glioblastoma subtypes in some populations compared to others, reflecting significantly better outcomes for patients^36,37^.

A further disparity in the findings between these two cohorts was identified in the study of epigenetic age in tumour tissues. Whilst the TCGA data revealed a relationship between epigenetic age acceleration and increasing tumour grade, this was not detected in the CGGA cohort, where the reverse was observed between astrocytomas of different grades. Direct comparisons of glioma subtypes between cohorts found a significant difference in only the LGA samples. Interestingly, this was the only glioma subtype where no significant difference was found in participant survival rates, suggesting that cohort-specific differences in epigenetic age acceleration alone are insufficient to explain differences in clinical outcome.

### Conclusions

Integrating DNA methylation profiles with transcriptomics and machine learning revealed distinct methylation signatures and biological pathways in glioblastomas from subjects of East Asian origin, compared to the non-Hispanic White glioma cohort, despite similar clinical classification. These findings suggest that molecular mechanisms driving glioma progression differ between ethnic groups and underscore the need to incorporate diverse populations into cancer genomics studies. Expanding the ancestral diversity of multi-omic datasets will be essential for improving molecular classification, biomarker discovery and the equitable implementation of precision oncology.

## Methods

### Datasets

Clinical and DNA methylation data was obtained from TCGA (TCGA-LGG, TCGA-GBM-27K and TCGA-GBM-450K) and CGGA (methyl_159, mRNA-array_301, mRNAseq_693 and mRNA sequencing (non-glioma as control))^11,38,39^. TCGA samples were selected from participants who had self-reported as White (non-Hispanic) ethnicity and only primary tumours from TCGA and CGGA were included in the study. All glioma samples were re-classified according to the WHO Classification of Tumours of the Central Nervous System, published in 2021^40^. In this study therefore, glioblastomas lack IDH mutations and astrocytomas possess an IDH mutation. The low-grade astrocytoma group includes astrocytomas of grade 2 only, and the high-grade astrocytoma group includes grades 3 and 4. Any participants present in both TCGA-GBM-27K and TCGA-GBM-450K were removed to avoid duplicate samples (*n* = 1).

Total participant numbers and patient characteristics are detailed in supplementary tables 1-3. Control data was obtained from the CGGA database (non-glioma brain tissue) and from the TCGA TARGET GTEx cohort via the USCS Xena browser^41,42^.

### DNA methylation analysis

For TCGA and CGGA datasets, DNA methylation data was collected using the Illumina Infinium HumanMethylation 27K or 450K BeadChip and is available as beta values. Beta values quantify the proportion of methylation cytosines and range from 0 to 1. Where required, beta values were converted to M-values by applying a logit transformation. Samples were removed where beta density profiles were not bimodal (n=15). CpG probes were removed which had >50% missing values in any dataset. Data imputation was performed on all TCGA participants using the 5-K Nearest Neighbour (KNN) algorithm from the impute R package. When combining datasets generated from different original cohorts, normalization was performed on M-values of all samples using Combat (surrogate variable analysis, sva R package). The participants from the TCGA dataset used in DNA methylation analysis included 110 glioblastoma samples for the analysis of 27K array data and 72 glioblastoma samples for 450K array data analysis.

Methylation-driven cancer genes (MDCGs) were identified using the MethylMix R package. This method identifies methylation drivers as those that are differentially methylated compared to controls, and that have a significant negative correlation with matched gene expression data. The CGGA datasets methyl_159 and mRNA-array_301 were used and included participants for which both sets of data were available (Supplementary Table 1). 11, 112 genes were included in the analysis, and where multiple CpG probes were available for a gene, the mean beta value was used.

MDCGs were researched for previous association to gliomas using PubMed, the OpenTargets platform, the Human Protein Atlas (Human Protein Atlas available from www.proteinatlas.org) and the DISEASES database (Jensen Lab)^43,44^.

Differential analysis of DNA methylation data was performed on normalised M-values using the limma R package, with empirical Bayes moderation and multiple testing correction using the Benjamini-Hochberg procedure. Adjusted p-values were used to identify significant changes in methylation of CpG probes. Enrichment analysis of gene ontology (GO) terms was performed using the methylglm function of the methylGSA Bioconductor R package. Statistical significance was determined using a logistic regression model in methylglm, with Benjamini-Hochberg correction for multiple testing.

Heatmaps of DNA methylation data were generated using the Pheatmap R package and beta density plots were created using Minfi R package. To assess molecular subtyping according to the DNA methylation of the GLDN gene, an unsupervised Gaussian mixture model was fitted using the Mclust R package and the relationship to actual glioma subtypes was represented using an alluvial diagram created using the ggalluvial R package.

### Transcriptomics analysis

The ‘mRNA sequencing (non-glioma as control)’ and ‘mRNAseq_693’ CGGA datasets were used for transcriptomics analysis of MDCGs (Supplementary Table 2). Participants present in both CGGA gene expression datasets (mRNA-array_301 and mRNAseq_693) were removed to create an independent validation set. Differential gene expression analysis was performed using DESeq2 on raw count data, and p-values corrected for multiple testing using the Benjamini-Hochberg procedure. Plots were created using the EnhancedVolcano and ggplot2 R packages. Correlation analysis between methylation and gene expression data was conducted using Pearson’s correlation analysis.

### Supervised machine learning model

For feature selection, differential analysis of DNA methylation data was performed (as described above) on each pair of glioma subtypes in the TCGA dataset. The significant probes with the highest log fold changes were obtained after this analysis and 265 unique CpG sites were selected as features. The random forest (RF) machine learning algorithm was applied to normalised M-values using the caret R package (CV=15, ntree=1000, train:test split = 70:30). Overall accuracy was calculated as: *true positives + true negatives / total observations*. Heatmaps were created from beta values (derived from normalized M-values using an inverse logit function), using the pheatmap R package, both with and without unsupervised hierarchical clustering of samples.

### Epigenetic age analysis and survival analysis

Epigenetic age was calculated using the Horvath (2013), PhenoAge and AltumAge algorithms in R (EpigeneticAgePipeline package: Horvath and PhenoAge) and Python (Pyaging package: AltumAge). Epigenetic Age Acceleration (EAA) was determined using the residual of difference between chronological age and epigenetic age. Kaplan-Meier survival analysis was performed using clinical data records of overall survival length and current status at time of data collection, using the ggsurvfit, Survival and Survminer R packages. Statistical differences were determined using the log-rank test and the Wald test.

### Statistical methods

Data analysis was conducted using R (version 4.5.1) and Python (version 3.9.21). Differences between groups were assessed, first using the F-test to determine variance, then with two-tailed *t*-tests. Significantly differentially expressed/methylated genes were identified using the Wald test and t-tests with empirical Bayes moderation, respectively. The Benjamini-Hochberg method was used to correct for multiple testing and *p*-values/adjusted *p*-values of <0.05 were considered statistically significant.

## Declarations

### Competing interests

The authors declare no competing interests.

### Ethics approval and consent to participate and publish

This study involved only the use of publicly available, de-identified data from The Cancer Genome Atlas and Chinese Glioma Genome Atlas.

### Availability of data and materials

The data supporting the results of this study are available upon reasonable request to the corresponding author.

### Author contributions

All authors contributed to the study conception and design. Data analysis and manuscript drafting was performed by Louise Newman. All authors commented on previous versions of the manuscript and approved the final manuscript.

### Funding

This work was financially supported by a Wellcome Trust ASPIRE award given to VWTC and ASO.

## Data Availability

All data produced in the present study are available upon reasonable request to the authors

## Acknowledgements

None.

## Supplementary information

**Supplementary Table 1.**
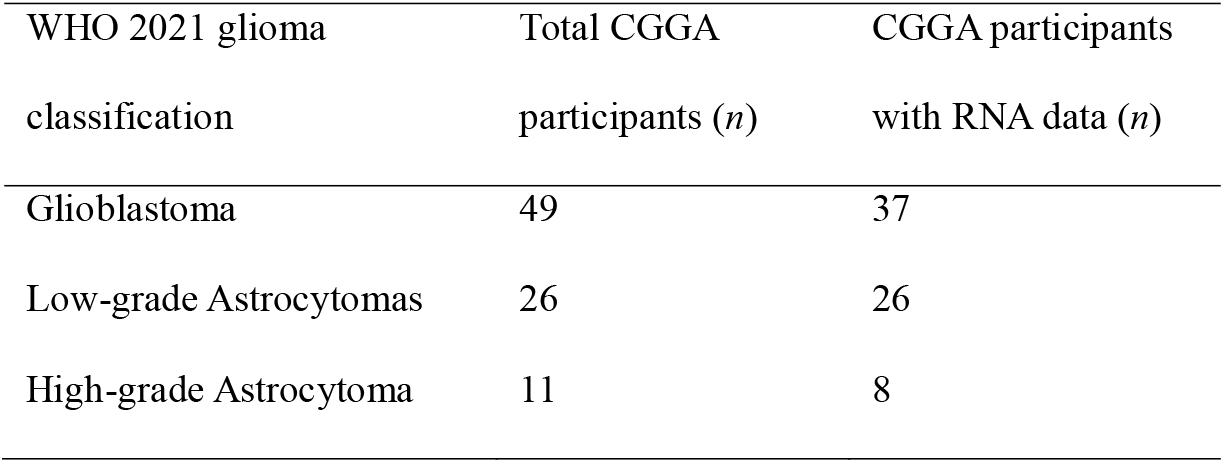
Participants from CGGA database for MethylMix analysis. Total participant numbers for each CGGA glioma subtype and the numbers for which there is also mRNA array data available for MethylMix analysis. Eight control samples were available for the analyis.

| WHO 2021 glioma classification | Total CGGA participants (n) | CGGA participants with RNA data (n) |
| --- | --- | --- |
| Glioblastoma | 49 | 37 |
| Low-grade Astrocytomas | 26 | 26 |
| High-grade Astrocytoma | 11 | 8 |

**Supplementary Table 2.** Participants from CGGA and TCGA database for transcriptomics analysis. Total participant numbers for each CGGA and TCGA (Non-Hispanic White only) glioma subtype and glioma subtype and comes from the ‘mRNAseq693’ dataset.

| WHO 2021 glioma classification | Total CGGA participants (n) | Total TCGA participants (n) |
| --- | --- | --- |
| Control | 20 | 1142 |
| Glioblastoma | 170 | 108 |
| Low-grade Astrocytomas | 51 | 38 |
| High-grade Astrocytoma | 71 | 72 |

**Supplementary Table 3.** Patient characteristics of participants included in DNA methylation analysis.

| Characteristic | CGGA Low-Grade Astrocytoma | CGGA High-Grade Astrocytoma | CGGA Glioblastoma | TCGA Low-Grade Astrocytoma | TCGA High-Grade Astrocytoma | TCGA Glioblastoma |
| --- | --- | --- | --- | --- | --- | --- |
| Total (n) | 26 | 11 | 49 | 38 | 73 | 110 |
| Age (median years, inter-quartile range) | 36.0 (38.8-30.5) | 43.0 (40.5-56.0) | 45.0 (35.0-56.0) | 36.0 (30.3-41.0) | 37.0 (33.0-46.0) | 60.0 (50.25-67.0) |
| Male (n, %) | 15 (57.7) | 7 (63.6) | 29 (59.2) | 20 (52.6) | 43 (58.9) | 64 (58.20) |
| Female (n, %) | 11 (42.3) | 4 (36.4) | 20 (40.8) | 18 (47.4) | 30 (41.1) | 46 (41.8) |

**Supplementary Table 4.**
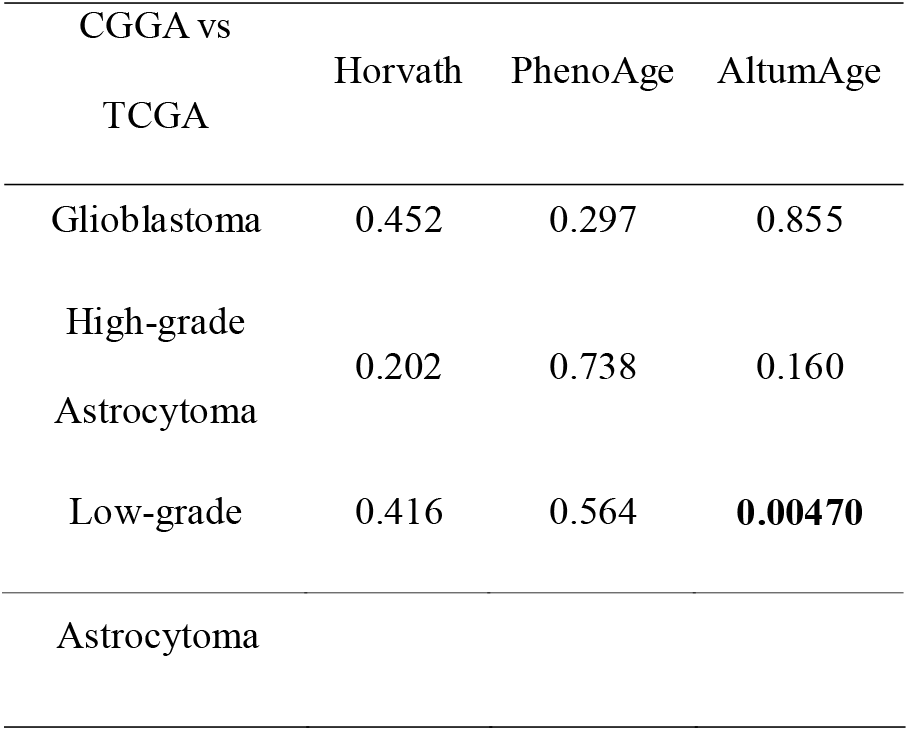
Epigenetic age acceleration of glioma types between CGGA and TCGA cohorts: Table of p-values, bold p-value indicates a statistically significant result.

**Supplementary Table 5.**
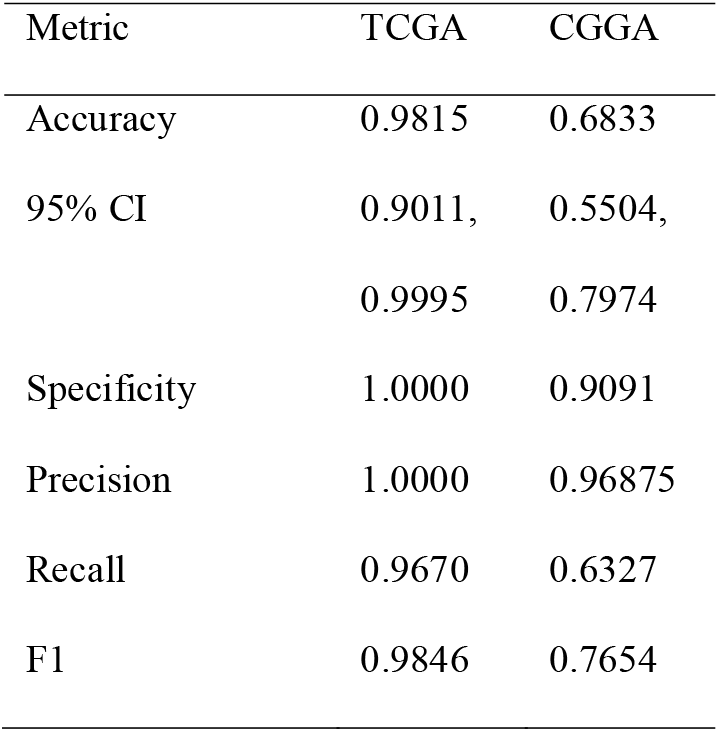
Performance metrics for supervised machine learning random forest models: Glioblastoma is the positive class, CI = confidence interval.

**Supplementary Table 6.**
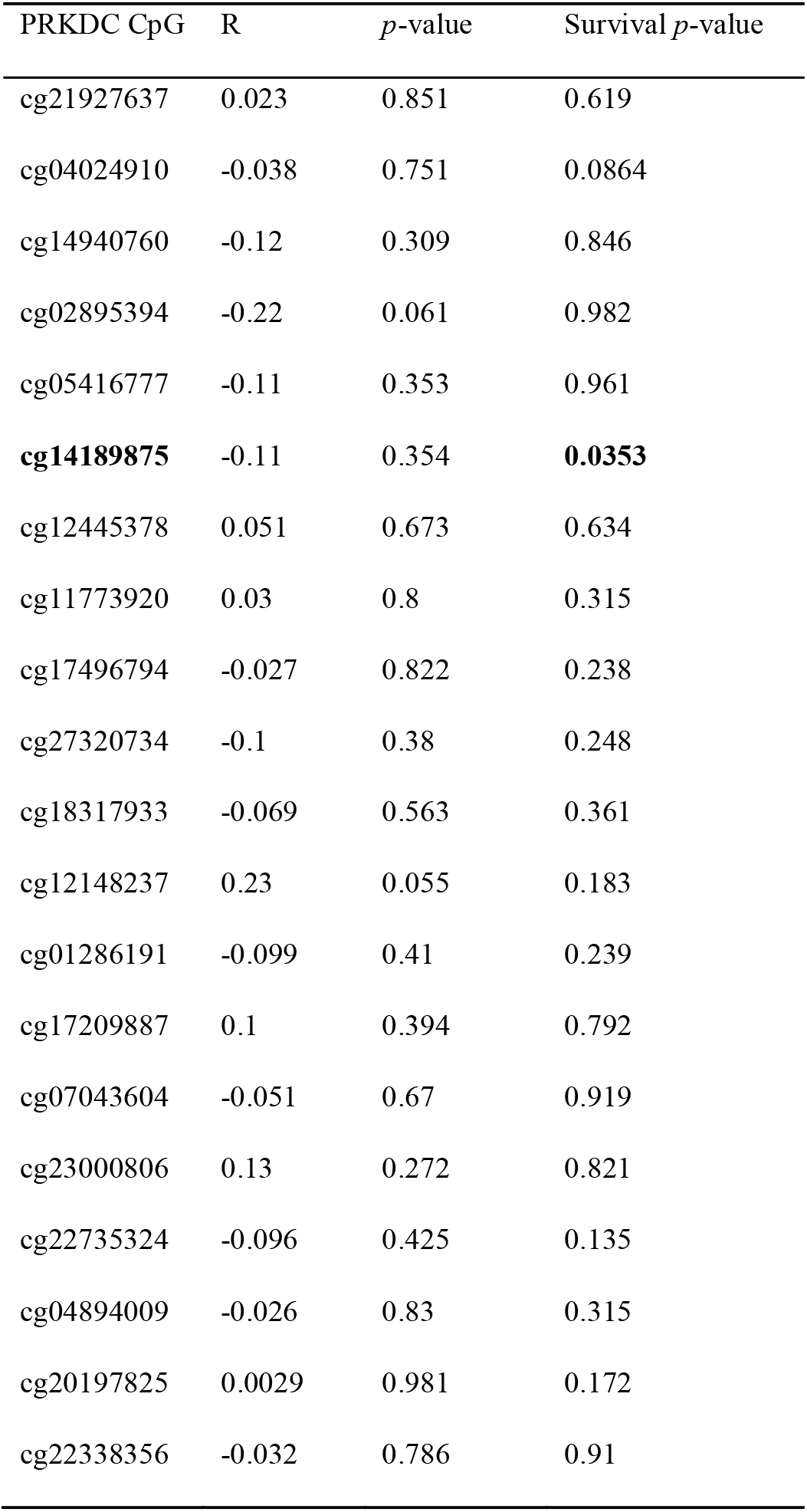
Correlation analysis and survival analysis of each PRKDC 450K array probe beta values compared with PRKDC gene expression in the glioblastoma samples from the TCGA dataset. Columns ‘R’ and ‘p-value’ describe the correlation analysis using Pearson’s correlation analysis. Column ‘Survival p-value’ represents the statistical significance between survival of above- and below-median PRDKC methylation groups. Statistical significance was determined using the Wald test. Values of statistical significance and the corresponding CpG probes are shown in bold.

## Supplementary Figures

**Supplementary Figure 1.**
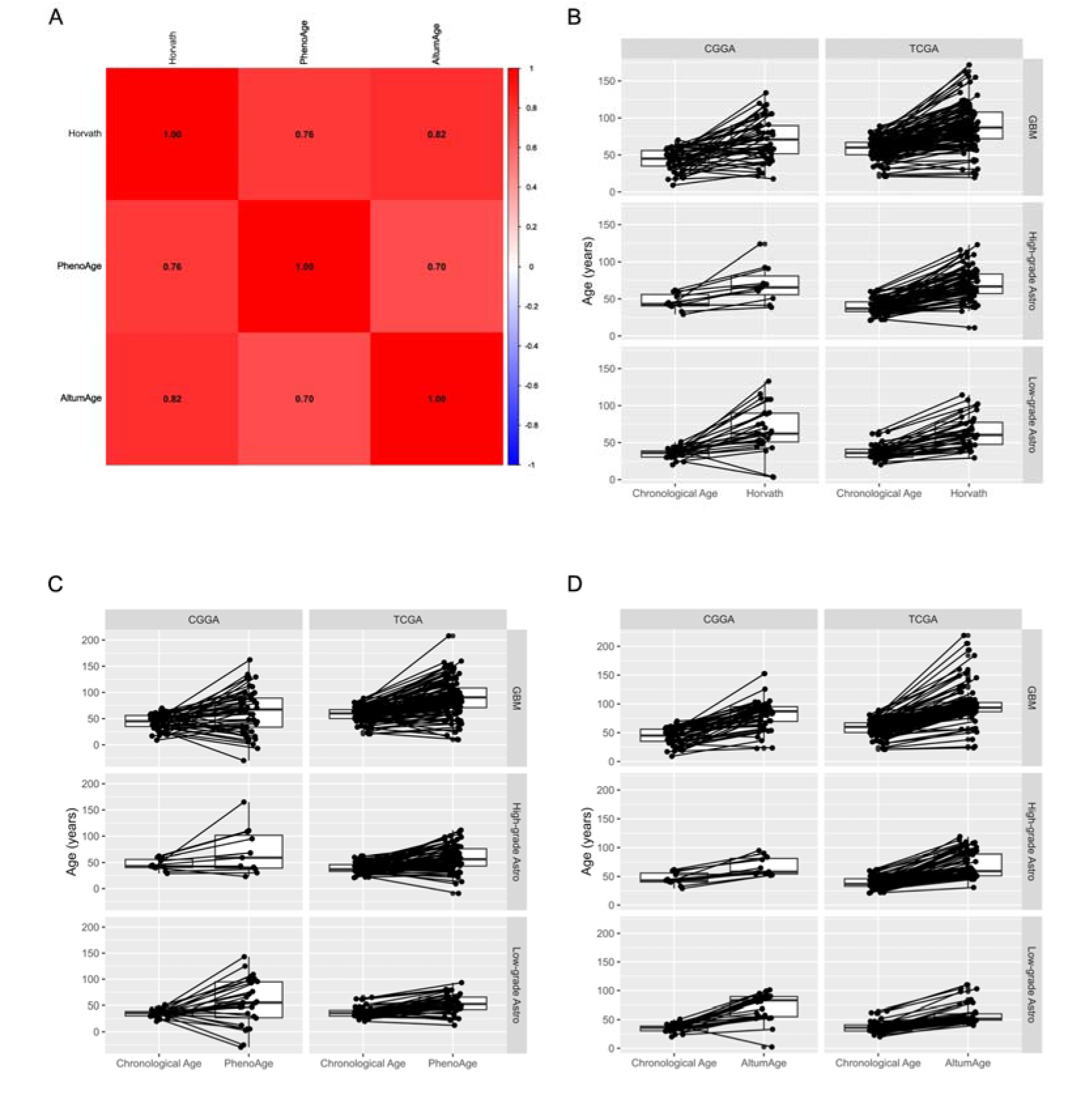
A. Correlation matrix of epigenetic age calculated using Horvath (2013), PhenoAge and AltumAge algorithms, on all glioma types from both cohorts. B-D. Box and whisker plots showing chronological age (left) and epigenetic age (right) measured in years, as estimated using epigenetic clock algorithms Horvath (B), PhenoAge (C) and AltumAge (D). Samples were separated by glioma type and cohort. Connections represent the direct age difference for each individual participant.

**Supplementary Figure 2.**
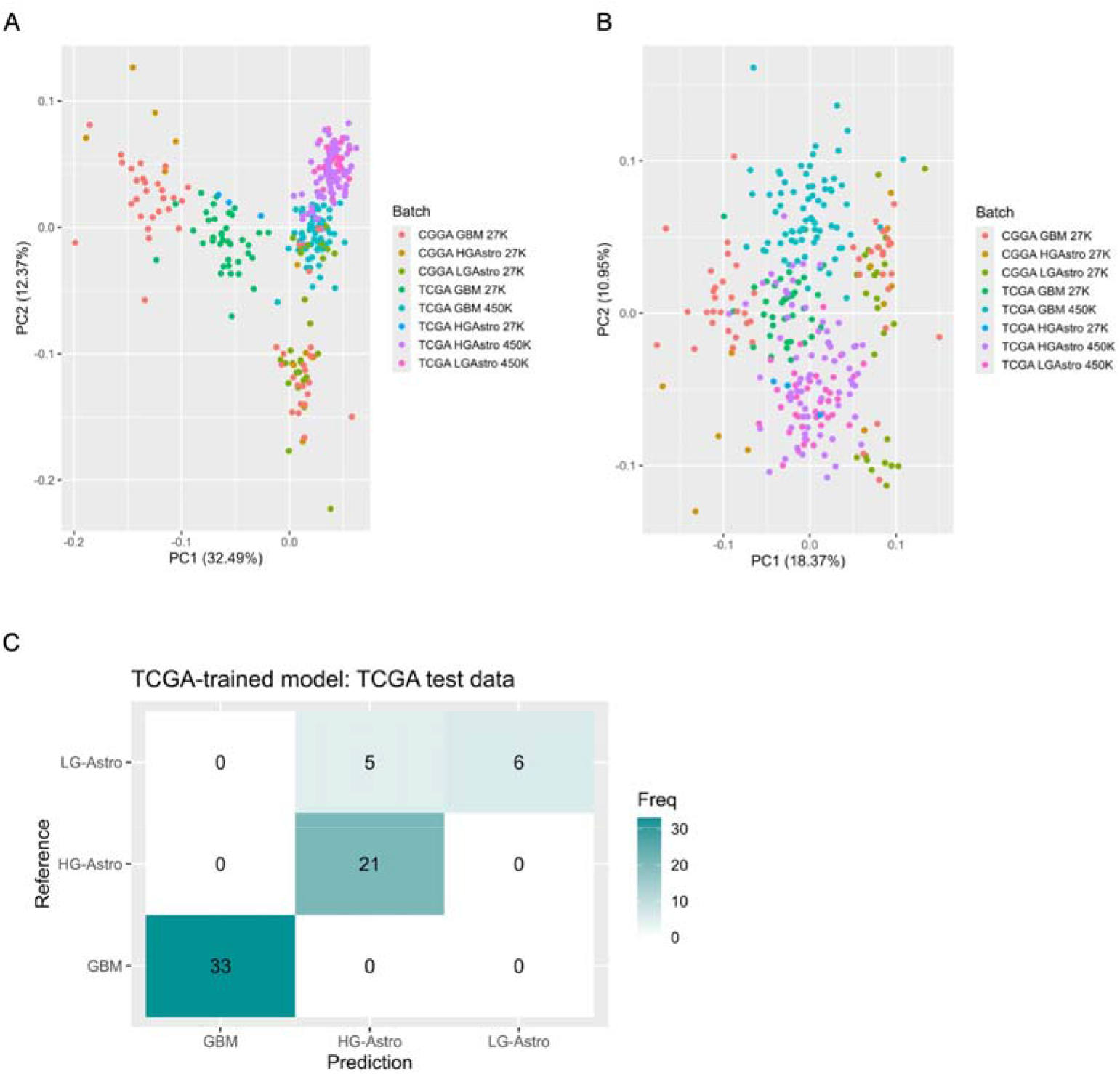
A-B. PCA plots before (A) and after (B) batch correction of M-values using ComBat. Batch corrected M-values were used for machine learning to reduce impact of technical differences between different experimental assays. C. Confusion matrix showing classification performance of Random Forest (RF) modelling on TCGA test data using batch corrected M-values of 265 CpG probes. Model was trained on a training set of TCGA data, classifying glioblastomas (GBM), high-grade astrocytomas (HG-Astro) and low-grade astrocytomas (LG-Astro).

**Supplementary Figure 3.**
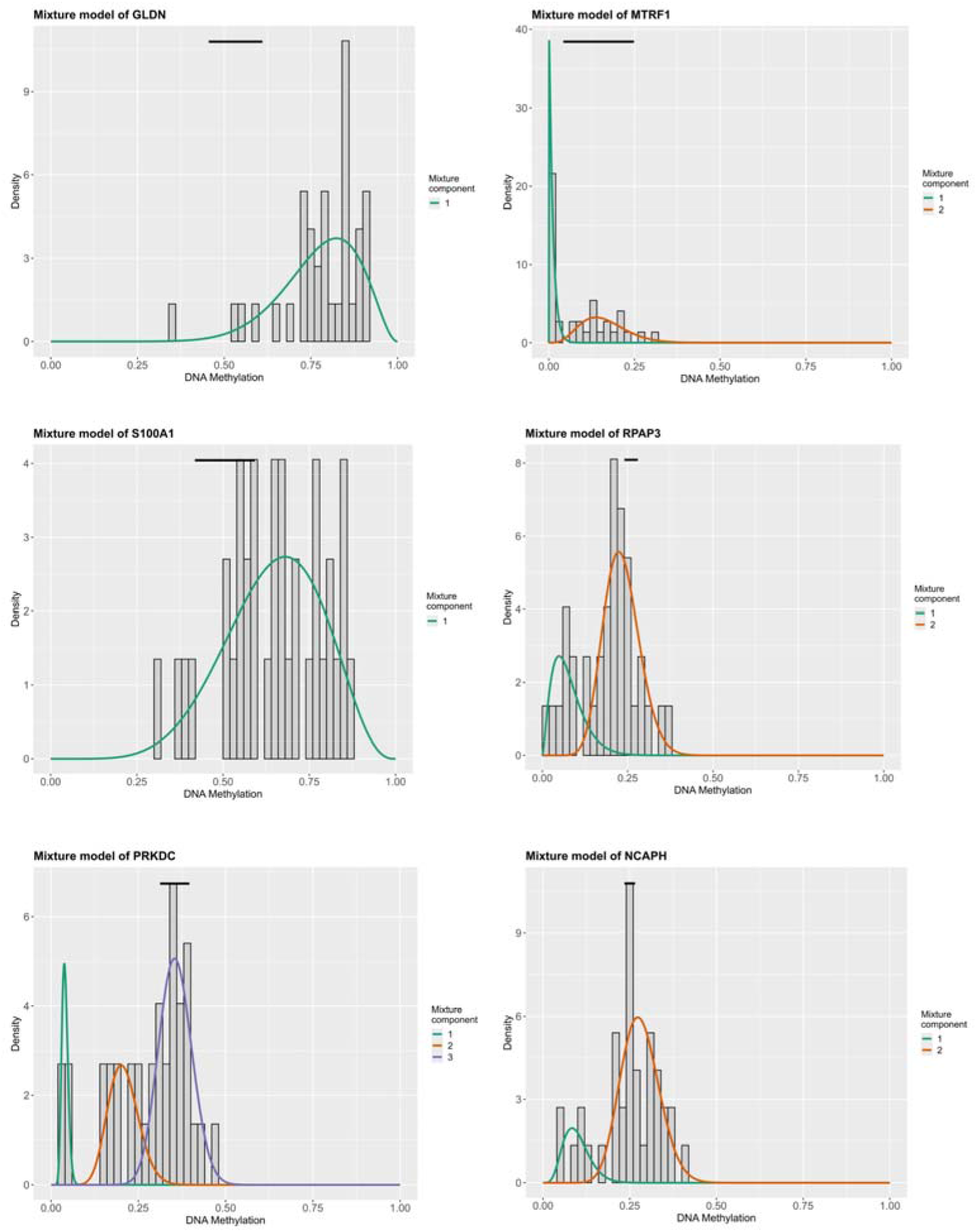
MethylMix Mixture models of the six MDCG identified in the CGGA glioblastoma samples. Density plots of the DNA methylation beta values in tumour samples. Black line indicates beta values of the control samples.

**Supplementary Figure 4.**
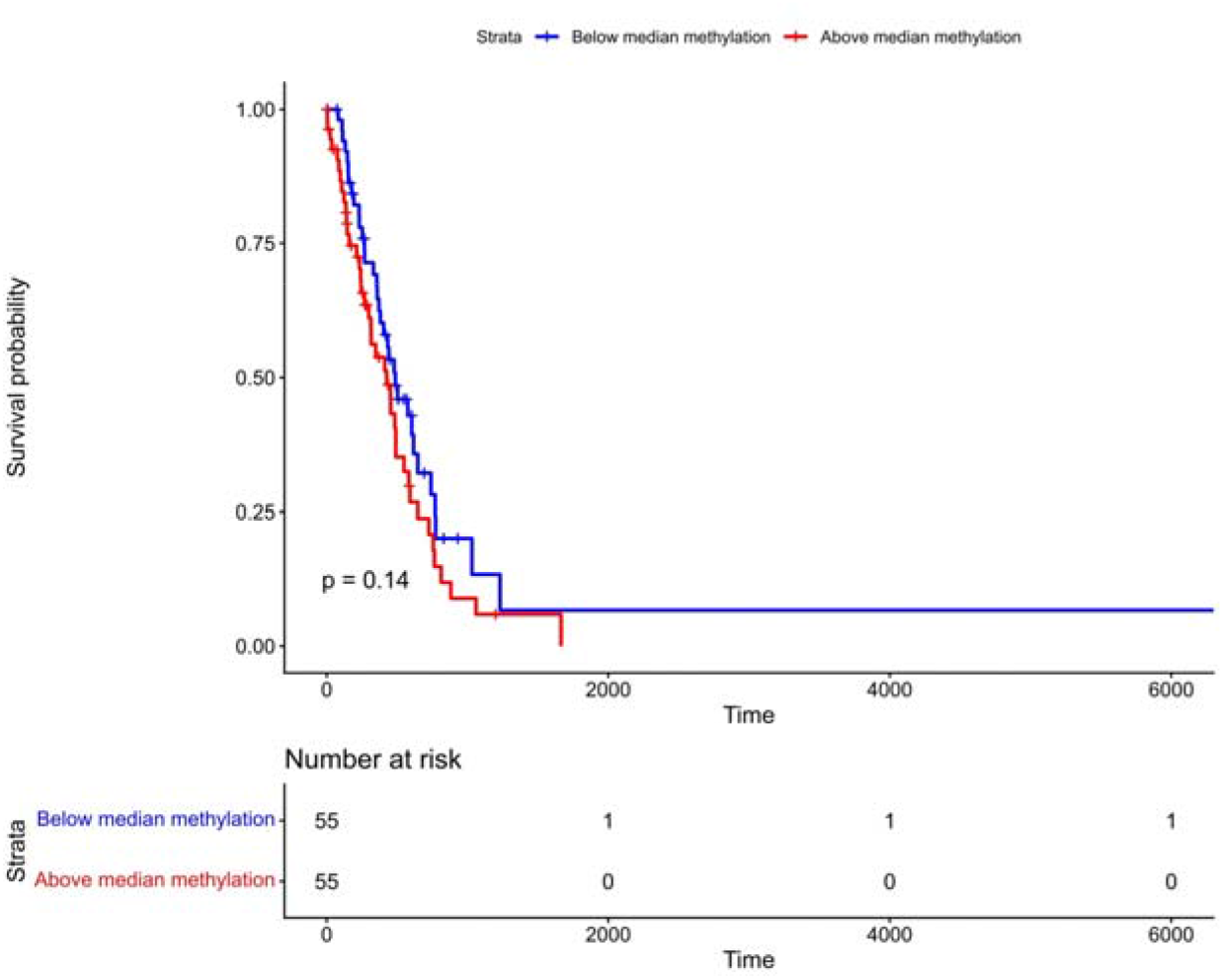
Kaplan-Meier survival analysis in glioblastoma samples from the TCGA cohort separated by PRKDC DNA methylation levels (cg23109897). Samples with DNA methylation beta values higher than the median are labelled in red, and those with beta values below the median are labelled in blue. Statistical significance was determined using the log-rank test.

**Supplementary Figure 5.**
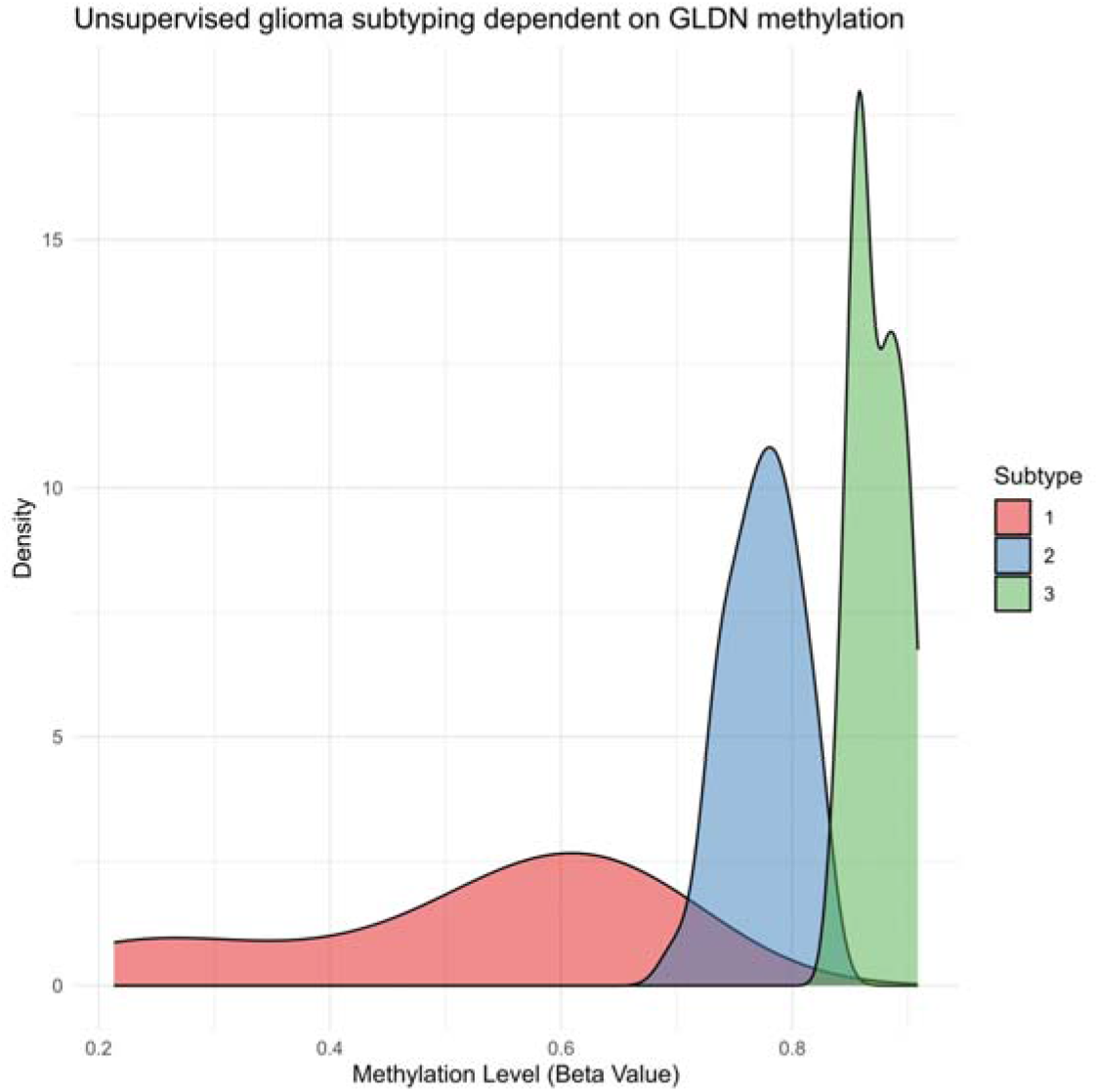
Beta density plot representing the glioma subtypes according to an unsupervised Gaussian mixture model using GLDN methylation data.

## Notes

### Competing Interest Statement

The authors have declared no competing interest.

### Author Declarations

The study used only openly available human data that were originally located at: Chinese Glioma Genome Atlas and The Cancer Genome Atlas

### Summary of Updates

This version has been updated to correct a calculation error in the data for Fig2b. The percentage calculation has been rectified (now 36.7%, previously 58.1%) and the corresponding text has been amended accordingly. The overall study conclusions remain unchanged.

